# Real-World Association Between Elevated Lipoprotein(a) and Cerebrovascular and Cardiovascular Outcomes: A Federated EHR Cohort Study

**DOI:** 10.64898/2026.04.30.26352189

**Authors:** Meet Popatbhai Kachhadia, Piyush Puri, Juber D. Shaikh, Marc A. Swerdloff

## Abstract

**Background:** Elevated lipoprotein(a) [Lp(a)] is causally implicated in atherosclerotic cardiovascular disease, but its prospective association with incident ischemic stroke in real-world clinical populations remains incompletely characterized. Observational data are complicated by testing-indication bias and aggressive preventive management in identified high-Lp(a) patients.

**Methods:** We conducted a retrospective cohort study using the TriNetX US Collaborative Network (66–67 healthcare organizations). Adults with Lp(a) measurements (LOINC 10835-7; January 2015–December 2025) were categorized as C-HIGH (Lp(a) ≥50 mg/dL) or C-LOW (<50 mg/dL) and balanced by 1:1 propensity score matching on cardiovascular diagnoses, medications, tobacco history, and four laboratory variables. Pre-specified analyses included an alternative cutoff (≥30 mg/dL), dose-response evaluation across three strata, and a 2-year landmark analysis.

**Results:** After expanded propensity score matching, the primary analysis included 97,882 matched adults (48,941 per arm). Incident ischemic stroke/TIA occurred in 2.45% vs 2.69% (Cox HR 0.956, 95% CI 0.878–1.041; log-rank p=0.299). The Lp(a) ≥30 mg/dL sensitivity analysis (121,076 matched) yielded HR 0.947 (0.878–1.023; p=0.168). Dose-response analysis showed no significant association at any stratum. A 2-year landmark analysis confirmed a null late-period effect (HR 1.071, 0.905–1.268; p=0.427). A post-hoc composite (stroke/TIA, cardiac arrest, heart failure) was null (HR 0.979; p=0.512). A broader MACE-like composite including MI yielded a nominally significant Cox HR of 1.058 (1.004– 1.116; p=0.034), with entirely null crude estimates, attributable to differential follow-up time.

**Conclusions:** Elevated Lp(a) was not associated with incident ischemic stroke or TIA across multiple thresholds and follow-up windows in this large federated cohort. These real-world findings are consistent with testing-indication bias and treatment attenuation in clinically identified high-Lp(a) populations.

## INTRODUCTION

Lipoprotein(a) [Lp(a)] is a genetically determined, atherogenic lipoprotein composed of apolipoprotein B-100 covalently bound to apolipoprotein(a). Plasma concentrations are approximately 70–90% heritable, relatively stable across a lifetime, and largely unresponsive to conventional lipid-lowering therapy.^1^

Mendelian randomization (MR) studies have provided compelling evidence for a causal role of Lp(a) in coronary artery disease and aortic valve stenosis. For ischemic stroke, MR studies report modestly elevated risk at higher Lp(a) concentrations, predominantly driven by large artery atherosclerotic mechanisms; a 2026 MR study published in this journal found a genetically predicted 100 nmol/L increase in Lp(a) was associated with large artery atherosclerotic stroke (OR 1.23, 95% CI 1.14–1.33) but not small vessel stroke (OR 0.98).^2,3^

Despite this genetic evidence, translating Lp(a)’s causal stroke risk to real-world clinical populations is not straightforward. Patients who receive clinical Lp(a) measurement are systematically different from the general population: they typically undergo testing due to premature or recurrent cardiovascular events, family history, or elevated calculated risk. Once identified, high-Lp(a) patients often receive intensified preventive management, introducing testing-indication and treatment-by-indication biases that may attenuate the observed Lp(a)–stroke association in observational data.

Novel Lp(a)-lowering agents, including the small interfering RNA olpasiran and the antisense oligonucleotide pelacarsen, have demonstrated 80–98% reductions in Lp(a) in phase 2 randomized trials, with pivotal phase 3 cardiovascular outcome trials ongoing.^5,6^ Understanding the real-world association between Lp(a) and vascular outcomes across the full concentration distribution is directly relevant to interpreting future trial data and positioning these agents for cerebrovascular indications. Prior observational studies of Lp(a) and stroke have been limited by relatively small sample sizes, single-center designs, incomplete covariate adjustment, and absence of dose-response characterization. We used the TriNetX US Collaborative Network, spanning 66–67 healthcare organizations, to conduct a comprehensive propensity-matched cohort study incorporating primary, alternative-cutoff, dose-response, and landmark analyses.

## METHODS

### Data Source and Study Design

We used the TriNetX Research Network US Collaborative Network (TriNetX, LLC; Cambridge, MA), a federated health research network providing standardized access to de-identified electronic medical records from 66–67 healthcare organizations across the United States. Data elements include diagnoses (ICD-10-CM), procedures (CPT/ICD-10-PCS), medications (RxNorm), laboratory values (LOINC), and vital signs. The observation period spanned January 1, 2015, through December 31, 2025. This study used de-identified data compliant with the HIPAA Privacy Rule; institutional review board oversight is not required under 45 CFR 46.101(b)(4).

We conducted a retrospective, new-user, active-comparator cohort study with 1:1 propensity score matching. Reporting follows the STROBE (Strengthening the Reporting of Observational Studies in Epidemiology)^8^ and RECORD-PE guidelines for studies using routinely collected health data.

### Cohort Definition and Eligibility

The study population comprised adults aged ≥18 years with at least one serum or plasma Lp(a) measurement (LOINC 10835-7; values 0–250 mg/dL) during the observation period. The index date was the date of the first qualifying Lp(a) measurement. Patients whose index event occurred more than 20 years before the analysis date were excluded.

Two primary exposure cohorts were defined: C-HIGH (Lp(a) ≥50 mg/dL) and C-LOW (Lp(a) <50 mg/dL). The ≥50 mg/dL threshold represents the international consensus high-risk cutoff endorsed by the European Atherosclerosis Society.^4^ A pre-specified alternative-cutoff sensitivity cohort used Lp(a) ≥30 mg/dL (C-HIGH-30) versus Lp(a) <30 mg/dL (C-LOW-30).

Exclusion criteria included prior intracranial hemorrhage (I60–I62), primary or secondary brain neoplasm (C71, D33, D43), necrotizing vasculopathies (M31), demyelinating disease (G36–G37), sickle cell disorders (D57), and antiphospholipid syndrome (D68.61).

### Primary Outcome

The primary outcome was incident ischemic stroke or TIA, defined by the first occurrence of ICD-10-CM codes I63 (cerebral infarction) or G45 (transient cerebral ischemic attacks) after the index date. Follow-up began 1 day post-index. Patients with any stroke/TIA diagnosis prior to the index date were excluded from the primary outcome analysis to ensure incident event capture.

### Propensity Score Matching

Propensity scores were estimated using logistic regression with the following pre-specified covariate blocks: (1) endocrine, nutritional, and metabolic diseases (E00–E89); (2) other forms of heart disease (I30–I52); (3) ischemic heart disease (I20–I25); (4) cerebrovascular disease (I60–I69); (5) hypertensive diseases (I10–I1A); (6) cardiovascular medications (CV000); (7) nicotine dependence (F17); (8) tobacco use (Z72.0); (9) personal history of nicotine dependence (Z87.891); (10) LDL cholesterol (LOINC 9002); (11) HbA1c (LOINC 9037); (12) estimated glomerular filtration rate by CKD-EPI 2021 (LOINC 98979-8); and (13) body mass index (LOINC 9083). Demographic covariates (age, sex, race) were included automatically by the TriNetX platform.

Matching was performed using 1:1 greedy nearest-neighbor matching without replacement. Post-match covariate balance was assessed using standardized mean differences (SMD); |SMD| <0.1 was considered acceptable. An SMD of 0.113 was observed for LDL-C in the primary analysis and is reported transparently as a limitation.

### Statistical Analysis

The primary effect estimate was the hazard ratio (HR) from a Cox proportional hazards model, with Kaplan-Meier survival curves compared by the log-rank test. The proportional hazards (PH) assumption was tested using a chi-square test based on weighted Schoenfeld residuals. Crude measures of association (risk difference, risk ratio, odds ratio) were calculated but interpreted cautiously given differential follow-up time between cohorts.

Pre-specified sensitivity and additional analyses included: (S4) alternative cutoff Lp(a) ≥30 vs <30 mg/dL; (S5) dose-response analysis evaluating three Lp(a) strata matched separately against C-LOW-30; (S6) a 2-year landmark analysis restricting outcome ascertainment to days 730–1,825 post-index; and post-hoc composite endpoint analyses (S7, S8) described in detail in the Results and Supplementary Material. Quantitative bias analysis used the E-value framework.7 Statistical significance was set at two-sided p<0.05. All analyses were conducted within the TriNetX federated platform (Study ID: 365036).

## RESULTS

### Study Population and Cohort Construction

In the primary analysis, the TriNetX US Collaborative Network contributed 48,409 C-HIGH patients (from 66 HCOs) and 81,422 C-LOW patients. After 1:1 expanded PSM, each cohort comprised 48,941 matched patients (97,882 total), reflecting matching retention of 97.7% in the C-HIGH arm (Figure 1).

**Figure 1.**
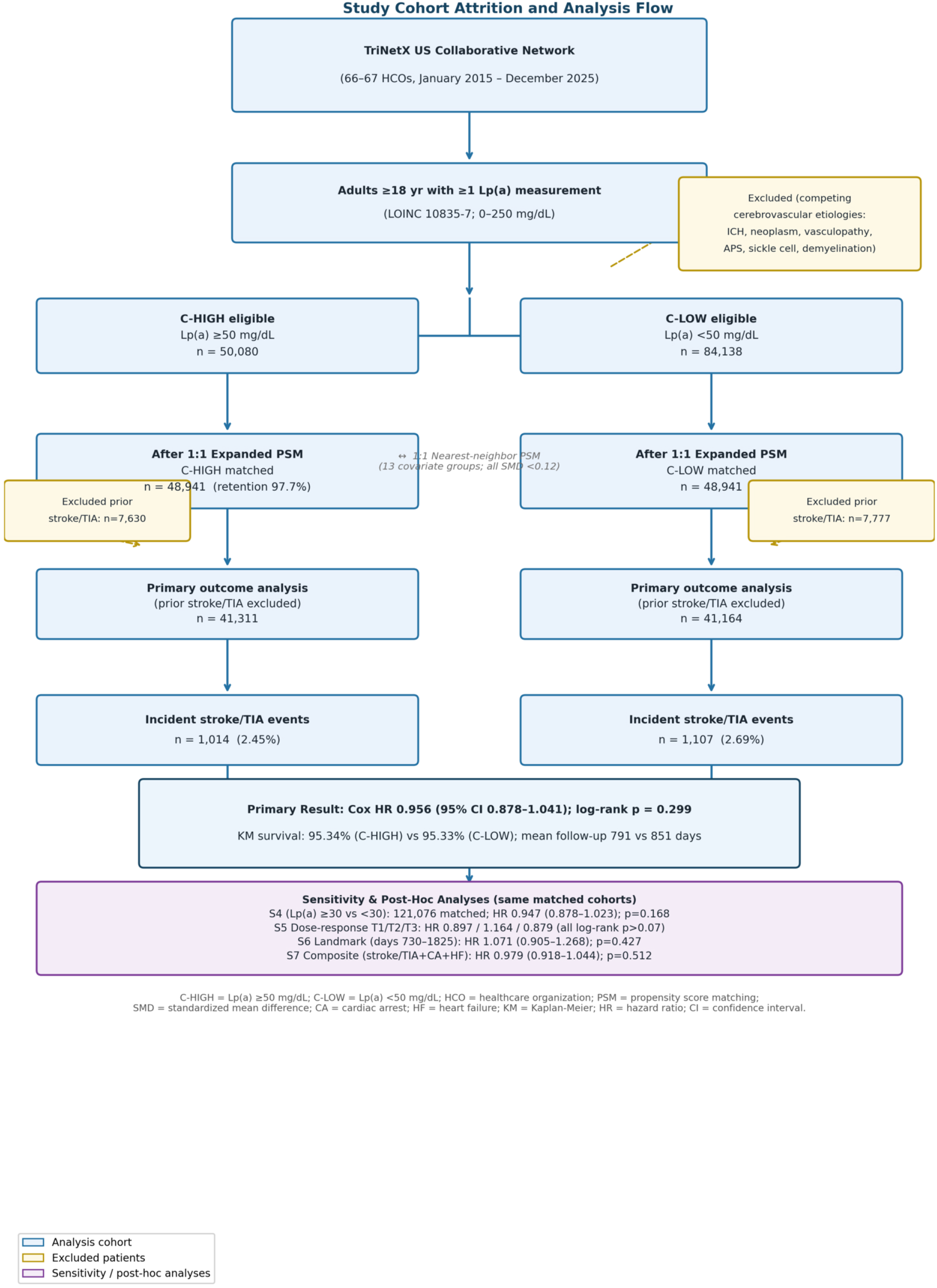
Study cohort attrition flowchart. Starting from the Lp(a)-measured base population in the TriNetX US Collaborative Network (66–67 HCOs), successive steps leading to the primary expanded-PSM cohorts and primary outcome analysis are shown. Primary analysis: C-HIGH 50,080 eligible → 48,941 matched; C-LOW 84,138 eligible → 48,941 matched. Numbers reflect the final analysis run (TriNetX Study ID 365036, April 2026). C-HIGH = Lp(a) ≥50 mg/dL; C-LOW = Lp(a) <50 mg/dL; HCO = healthcare organization; PSM = propensity score matching; SMD = standardized mean difference.

Expanded PSM achieved excellent covariate balance across diagnoses, medications, and tobacco history (all SMD <0.02), with residual imbalance limited to LDL cholesterol (post-match SMD 0.113) and eGFR (SMD 0.075) (Table 1). Mean follow-up after matching was 791.1 days (SD 674.7; median 570 days) in C-HIGH and 851.4 days (SD 697.5; median 657 days) in C-LOW.

**Table 1.**
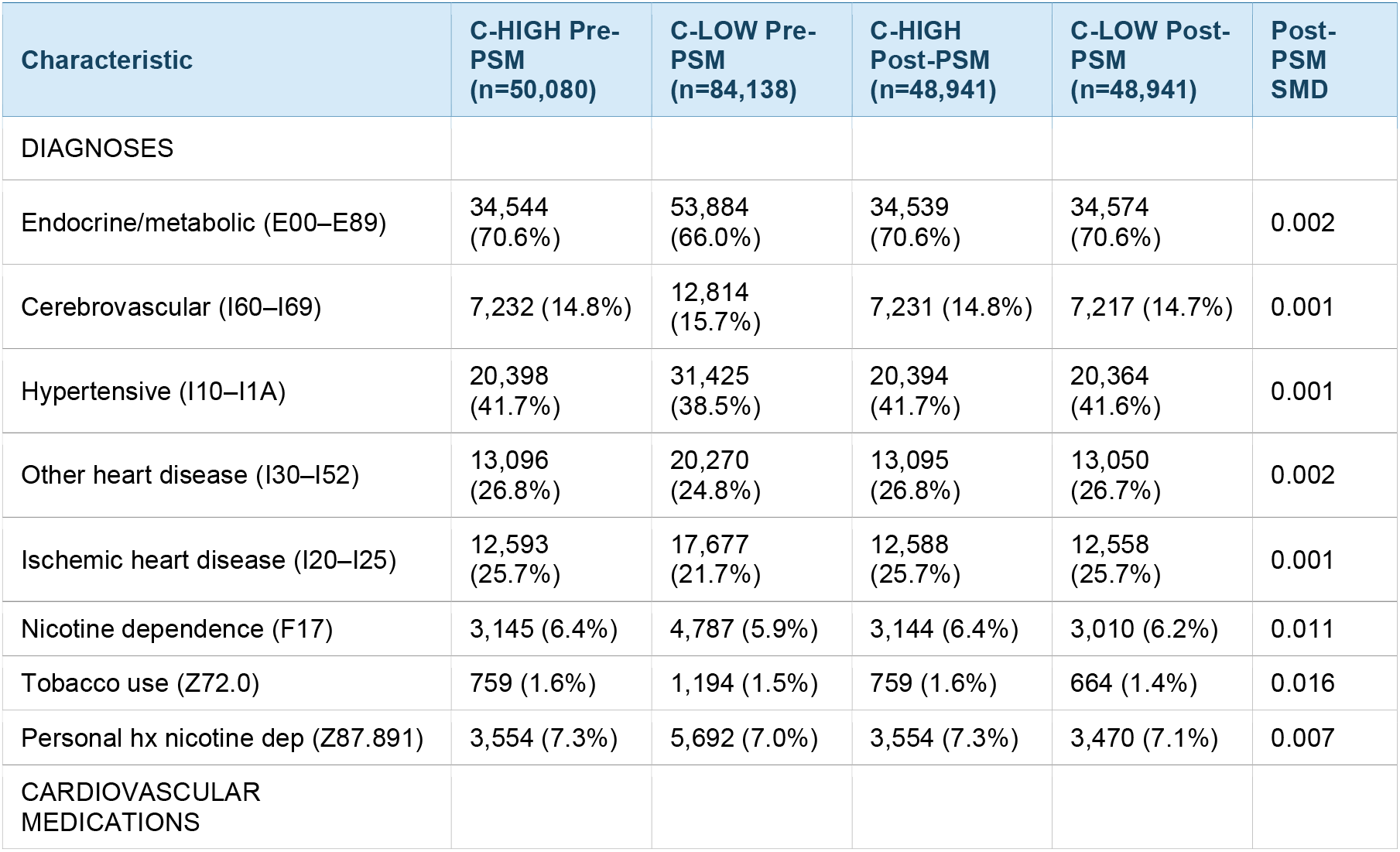
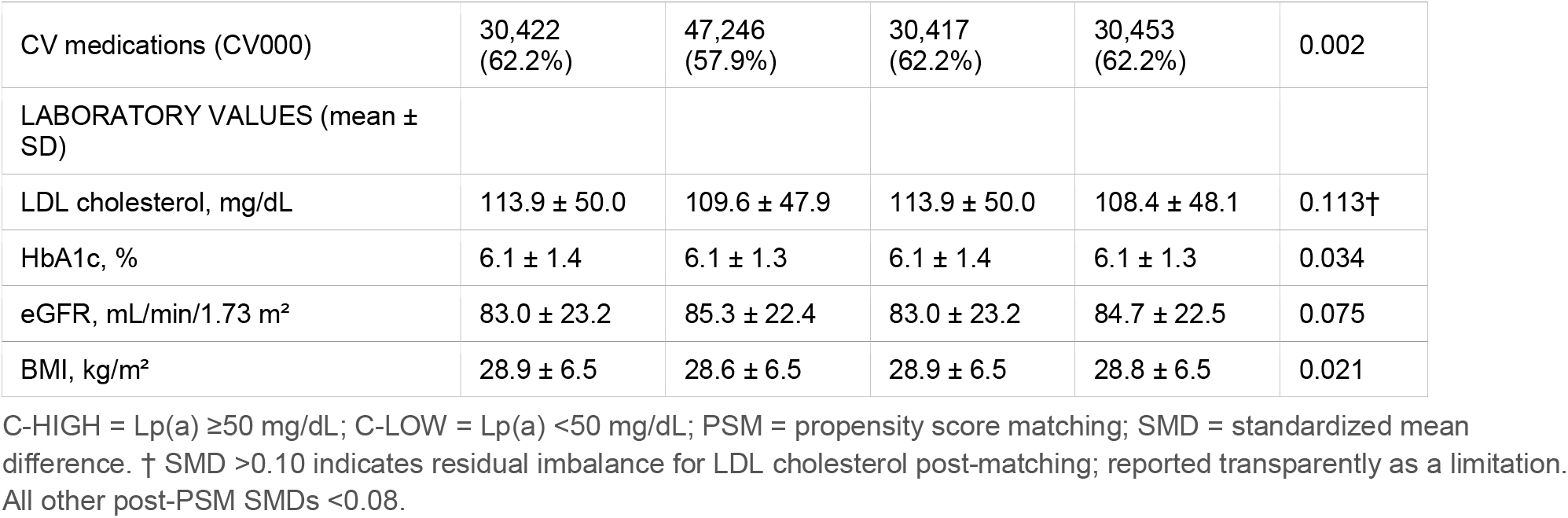
Baseline Characteristics Before and After Expanded Propensity Score Matching (Primary Analysis, Lp(a) ≥50 vs <50 mg/dL)

### Primary Outcome: Lp(a) ≥50 vs <50 mg/dL

After excluding patients with prior stroke or TIA (7,630 C-HIGH; 7,777 C-LOW), the incident-event analysis included 41,311 C-HIGH and 41,164 C-LOW patients. Incident stroke or TIA occurred in 1,014 of 41,311 (2.45%) C-HIGH patients and 1,107 of 41,164 (2.69%) C-LOW patients (Table 2).

**Table 2.** Primary Outcome Results: Incident Ischemic Stroke/TIA, Lp(a) ≥50 vs <50 mg/dL.

| Parameter | C-HIGH (Lp(a) ≥50 mg/dL) | C-LOW (Lp(a) <50 mg/dL) | p-value / Statistic |
| --- | --- | --- | --- |
| Matched patients (post-PSM) | 48,941 | 48,941 |  |
| Patients analyzed for primary outcome<sup>1</sup> | 41,311 | 41,164 |  |
| — Prior stroke/TIA excluded | 7,630 | 7,777 |  |
| Incident stroke or TIA events | 1,014 | 1,107 |  |
| Crude risk | 2.45% | 2.69% |  |
| Mean follow-up, days (SD) | 791.1 (674.7) | 851.4 (697.5) |  |
| Median follow-up, days | 570 | 657 |  |
| KM survival probability at end of window | 95.34% | 95.33% |  |
| Cox Hazard Ratio (95% CI)<sup>2</sup> | 0.956 (0.878–1.041) | Reference | p=0.299 |
| Log-rank chi-square (df=1) | — | 1.080 | p=0.299 |
| PH proportionality test (χ<sup>2</sup>, p) | — | χ<sup>2</sup>=4.322 | p=0.038<sup>3</sup> |
| Risk Difference (95% CI) | — | −0.002 (−0.005, −0.000) | p=0.033<sup>4</sup> |
| Risk Ratio (95% CI) | — | 0.913 (0.839–0.993) | p=0.033<sup>4</sup> |
| Odds Ratio (95% CI) | — | 0.911 (0.835–0.993) |  |
| E-value (point estimate / upper CI) | — | 1.18 / 1.0 |  |

<sup>1</sup> After exclusion of patients with prior stroke/TIA. <sup>2</sup> Cox HR is the pre-specified primary estimate. <sup>3</sup> PH assumption violated; log-rank test confirms null finding. <sup>4</sup> Crude proportion-based estimates; biased by differential follow-up time.

The Cox HR for incident stroke or TIA was 0.956 (95% CI 0.878–1.041). The log-rank chi-square was 1.080 (df=1; p=0.299). Kaplan-Meier survival probabilities at the end of the observation window were 95.34% (C-HIGH) and 95.33% (C-LOW). A mild violation of the proportional hazards assumption was noted (Schoenfeld residuals χ^2^=4.322; p=0.038); the log-rank statistic, which requires no proportionality assumption, independently confirms the null result. The E-value for the point estimate was 1.18.

### Sensitivity Analysis: Lp(a) ≥30 vs <30 mg/dL (S4)

After 1:1 expanded PSM, 60,538 patients were matched per arm (121,076 total). The Cox HR was 0.947 (95% CI 0.878–1.023; log-rank p=0.168). Concordance of the ≥50 and ≥30 analyses demonstrates threshold independence of the null finding.

### Dose-Response Analysis: Lp(a) Strata vs <30 mg/dL Reference (S5)

Across all three Lp(a) strata, no statistically significant increase in stroke/TIA risk was observed (Table 3). The dose-response pattern was non-monotonic (T1 HR 0.897, T2 HR 1.164, T3 HR 0.879), inconsistent with a concentration-dependent atherogenic gradient. Significant PH violations at T2 (p=0.006) and T3 (p=0.004) indicate time-varying hazard patterns; log-rank p-values are the primary inferential tests for these strata.

**Table 3.** Dose-Response Analysis: Incident Ischemic Stroke/TIA by Lp(a) Stratum vs <30 mg/dL Reference.

| Lp(a) Tier vs Ref (<30 mg/dL) | Matched N (per arm) | Analyzed C-Tier / C-LOW | Events C-Tier (%) | Events C-LOW (%) | HR (95% CI) | Log-rank p |
| --- | --- | --- | --- | --- | --- | --- |
| T1: 50–99 mg/dL | 25,027 | 20,850 / 20,817 | 504 (2.4%) | 583 (2.8%) | 0.897 (0.797–1.011) | 0.075 |
| T2: 100–149 mg/dL | 7,371 | 5,892 / 5,830 | 221 (3.8%) | 194 (3.3%) | 1.164 (0.960–1.412)<sup>†</sup> | 0.122 |
| T3: ≥150 mg/dL | 9,529 | 8,212 / 8,114 | 182 (2.2%) | 215 (2.6%) | 0.879 (0.722–1.071)<sup>†</sup> | 0.202 |

All strata matched separately against C-LOW-30 (<30 mg/dL) using the same expanded PSM specification. Prior stroke/TIA excluded from each analysis. <sup>†</sup> PH assumption violated at p<0.01; log-rank p-values are the primary inferential test.

### Landmark Analysis: Days 730–1,825 Post-Index (S6)

The Cox HR was 1.071 (95% CI 0.905–1.268; log-rank p=0.427). No PH violation was detected (p=0.934). These data confirm that the null primary result is not attributable to inadequate early follow-up.

### Post-Hoc Composite Analyses (S7 and S8)

In S7 (composite of ischemic stroke/TIA, cardiac arrest, and heart failure), incident composite events occurred in 4.89% (C-HIGH) versus 5.24% (C-LOW) of at-risk patients (Cox HR 0.979, 95% CI 0.918– 1.044; log-rank p=0.512). In S8 (broader MACE-like composite additionally incorporating MI, angina, cerebral/precerebral artery occlusion, intracranial hemorrhage, and cardiovascular events), incident events occurred in 8.8% vs 8.9% (Cox HR 1.058, 95% CI 1.004–1.116; p=0.034; PH test p=0.639). Crude risk difference was −0.000 (p=0.863), entirely null, reflecting differential follow-up time. The directional shift from S7 to S8 with addition of MI codes is consistent with Lp(a)’s stronger causal association with coronary versus cerebrovascular endpoints in MR studies. Full results are in Supplementary Tables S3–S4.

### Comprehensive Analysis Summary

Table 4 presents all primary and sensitivity analyses. Hazard ratios ranged from 0.879 to 1.164 across all pre-specified analyses; none achieved log-rank significance.

**Table 4.** Comprehensive Summary of All Primary and Sensitivity Analyses.

| Analysis | Matched N (per arm) | Events C-HIGH / C-LOW | Analyzed N C-HIGH / C-LOW | HR (95% CI) | Log-rank p | PH test p |
| --- | --- | --- | --- | --- | --- | --- |
| PRIMARY: Lp(a) ≥50 vs <50 mg/dL | 48,941 | 1,014 / 1,107 | 41,311 / 41,164 | 0.956 (0.878–1.041) | 0.299 | 0.038* |
| S4: Lp(a) ≥30 vs <30 mg/dL | 60,538 | 1,243 / 1,379 | 50,921 / 50,375 | 0.947 (0.878–1.023) | 0.168 | 0.054 |
| S5-T1: 50–99 vs <30 mg/dL | 25,027 | 504 / 583 | 20,850 / 20,817 | 0.897 (0.797–1.011) | 0.075 | 0.073 |
| S5-T2: 100–149 vs <30 mg/dL | 7,371 | 221 / 194 | 5,892 / 5,830 | 1.164 (0.960–1.412) | 0.122 | 0.006* |
| S5-T3: ≥150 vs <30 mg/dL | 9,529 | 182 / 215 | 8,212 / 8,114 | 0.879 (0.722–1.071) | 0.202 | 0.004* |
| S6 Landmark: days 730–1825 | 48,941 | 263 / 277 | 40,560 / 40,334 | 1.071 (0.905–1.268) | 0.427 | 0.934 |
| S7 Post-hoc composite<sup>†</sup> | 48,947 | 1,816 / 1,941 | 37,140 / 37,046 | 0.979 (0.918–1.044) | 0.512 | NR |
| S8 Broader MACE-like composite<sup>‡</sup> | 48,944 | 2,778 / 2,809 | 31,494 / 31,705 | 1.058 (1.004–1.116)* | 0.034 | 0.639 |

\* S8: Cox HR nominally significant (p=0.034) but crude RD –0.000 (p=0.863); discrepancy attributable to differential follow-up time. PH assumption satisfied (p=0.639). <sup>†</sup> S7: composite of ischemic stroke/TIA + cardiac arrest + HF; nonfatal MI not included. <sup>‡</sup> S8: broader MACE-like composite including MI, angina, precerebral artery events, intracranial hemorrhage, hypertensive heart disease, other circulatory disorders, and death; not pre-specified. NR = not reported.

## DISCUSSION

In this large propensity-matched federated cohort study of 97,882 adults across 66–67 US healthcare organizations, elevated Lp(a) assessed at four different thresholds and across three concentration strata was not associated with incident ischemic stroke or TIA over follow-up extending to 5 years. Hazard ratios from all six pre-specified analyses converged between 0.879 and 1.164, none achieving statistical significance on the log-rank test. A 2-year landmark analysis confirmed that the null finding persists in the late follow-up window, excluding delayed atherogenic effects over the study period.

These findings contrast with Mendelian randomization studies establishing a causal effect of genetically elevated Lp(a) on ischemic stroke, predominantly through large artery atherosclerotic mechanisms.^2,3^ The observed discrepancy is consistent with several non-exclusive explanations.

Testing-indication bias is likely the dominant mechanism. Patients who receive clinical Lp(a) measurement are systematically enriched for individuals with premature cardiovascular events, strong family histories, or high calculated risk — precisely those receiving the most aggressive pharmacologic management. The near-uniform cardiovascular medication exposure in both matched cohorts (62.2%) reflects this: high-Lp(a) patients in routine care are likely receiving statins, antiplatelets, and anticoagulation at rates comparable to or exceeding the lower-Lp(a) comparison group, effectively neutralizing the atherogenic signal.

The non-monotonic dose-response pattern deserves specific comment. At T2 (100–149 mg/dL), the point estimate was numerically elevated (HR 1.164), though non-significant (log-rank p=0.122) with a significant PH violation (p=0.006). At the highest Lp(a) concentrations (T3, ≥150 mg/dL), the HR was paradoxically 0.879, likely reflecting particularly aggressive preventive management in patients with extreme Lp(a) elevations. These non-monotonic findings are most parsimoniously explained by heterogeneous treatment-by-indication effects across concentration strata rather than genuine reversal of atherogenic risk.

The PH violations across multiple analyses indicate time-varying hazard patterns. The landmark analysis offers a partial solution: restricting to days 730–1,825, where no PH violation was detected (p=0.934), produced an HR of 1.071 (0.905–1.268) that is non-significant and consistent with the null, strengthening confidence in the null conclusion.

The discrepancy between significant crude risk ratios and the null Cox HR is explained entirely by differential follow-up time. C-LOW patients had approximately 8% longer mean follow-up (851 vs 791 days), providing more observation time to accumulate events. The Cox HR, conditioning on time-at-risk, is the methodologically correct primary estimate.

The post-hoc composite analyses add mechanistic resolution. The primary cerebrovascular finding remains robustly null in S7 (HR 0.979; log-rank p=0.512). The directional shift in S8 — from null toward elevated only when MI codes are added — is directly consistent with Lp(a)’s known stronger causal effect on coronary artery disease than on stroke in MR studies, and strengthens rather than undermines the paper’s central conclusion. The nominal Cox HR significance in S8 must be interpreted against the completely null crude risk difference (−0.000, p=0.863) and the differential follow-up artifact identified throughout this study. Additionally, ICD-10 codes I63/G45 do not discriminate by stroke mechanism; a recent MR study in this journal demonstrated a specific association with large artery atherosclerotic stroke (OR 1.23 per 100 nmol/L) but not small vessel stroke (OR 0.98),^2^ and this mechanistic dilution may further attenuate any signal in the overall stroke composite.

### Strengths and Limitations

Key strengths include the large sample size (>97,000 matched patients in the primary analysis, >121,000 in S4), multi-institutional federated design across 66–67 HCOs, expanded PSM with 13 covariate groups incorporating laboratory values, pre-specified analyses including dose-response and landmark designs, E-value bias quantification, and paired post-hoc composite endpoint analyses. Limitations are as follows. First and most critically, EHR-based Lp(a) measurement introduces testing-indication bias that PSM on observed covariates cannot fully resolve. Second, residual LDL-C imbalance (SMD 0.113–0.104) persists after expanded PSM; E-value analysis suggests it cannot materially alter the null conclusion. Third, significant PH violations in the primary analysis and dose-response strata indicate time-varying hazard patterns. Fourth, ICD-10-based stroke ascertainment has an estimated positive predictive value of approximately 80% compared with adjudicated outcomes. Fifth, TriNetX uses administrative date-of-last-fact as the censoring mechanism, potentially introducing informative censoring. Sixth, neither post-hoc composite endpoint maps directly to the 3-point MACE endpoints of Lp(a)HORIZON and OCEAN(a)-OUTCOMES. Seventh, the ICD-10 stroke code (I63) does not capture stroke etiology, preventing subtype-specific analysis.

## CONCLUSIONS

In this comprehensive propensity-matched federated cohort study, elevated Lp(a) was not associated with incident ischemic stroke or TIA across seven pre-specified and sensitivity analyses in 66–67 US healthcare organizations. Hazard ratios for the primary stroke endpoint were tightly null across all analyses (range 0.879–1.164; none log-rank significant), no dose-response gradient was evident, and a 2-year landmark analysis confirmed the absence of delayed atherogenic effects. These real-world findings are consistent with attenuation of Lp(a)’s cardiovascular signal driven by testing-indication bias and intensified preventive management. They do not contradict MR and trial evidence supporting Lp(a) as a causal cardiovascular risk factor. As novel Lp(a)-lowering agents progress through pivotal phase 3 cardiovascular outcomes trials, understanding the gap between genetic causal estimates and real-world observational associations is essential for contextualizing trial results and identifying populations most likely to benefit.

## CLINICAL PERSPECTIVE

### What Is New

In a propensity-matched federated EHR cohort of >97,000 adults across 66 US healthcare organizations, elevated Lp(a) (≥50 mg/dL) was not associated with incident ischemic stroke/TIA across seven independent analytic approaches. A post-hoc composite without MI (S7: stroke/TIA + cardiac arrest + HF) was null (HR 0.979; p=0.512). A second post-hoc composite additionally including MI and broader ischemic/circulatory events (S8) produced a nominally significant Cox HR of 1.058 (1.004–1.116; p=0.034), but crude risk estimates were entirely null, reflecting a differential follow-up artifact. The directional shift with MI inclusion is consistent with Lp(a)’s stronger MR-defined causal effect on coronary versus cerebrovascular endpoints.

### What Are the Clinical Implications

Null real-world associations are consistent with testing-indication bias: patients identified with high Lp(a) in routine clinical practice carry elevated cardiovascular risk, which in turn triggers aggressive preventive management that attenuates the expected atherogenic signal. These findings do not contradict MR or phase 2 trial evidence supporting Lp(a) as a causal cardiovascular risk factor and should not discourage Lp(a) screening or enrollment in ongoing phase 3 cardiovascular outcomes trials (Lp(a)HORIZON, OCEAN(a)-OUTCOMES).

## DISCLOSURES AND ACKNOWLEDGMENTS

### Data Availability

Analysis was conducted within the TriNetX federated platform (Study ID 365036). De-identified aggregate data are available upon reasonable request.

### IRB Statement

This study used de-identified federated data and does not meet the definition of human subjects research under 45 CFR 46.101(b)(4). No IRB approval was required.

### AI Disclosure

Artificial intelligence tools were used to assist in manuscript organization. All analytic decisions, data interpretation, and final manuscript content were reviewed and approved by the authors.

## AUTHOR CONTRIBUTIONS (CRediT)

Meet Popatbhai Kachhadia: Conceptualization, Data curation, Formal analysis, Writing — original draft, Project administration.

Piyush Puri: Conceptualization, Methodology, Formal analysis, Writing — review and editing, Supervision.

Juber D. Shaikh: Writing — review and editing.

Marc A. Swerdloff: Writing — review and editing, Supervision.

